# Oxygen extraction fraction brain charts for human lifespan and application for brain disorders

**DOI:** 10.64898/2026.06.02.26354684

**Authors:** Zixuan Lin, Xiang Fan, Tianyu Gao, Shuyue Wang, Yiwen Hong, Yifan Yan, Jianpeng Liu, Yuchuan Fu, Tao Hua, Yue Cai, Gaigai Lu, Ying Qi, Bing Yu, Zhizheng Zhuo, Jiani Wu, Dayong Ge, Qingyu Xu, Yizhe Hu, Chuhan Xiong, Weijia Liu, Runyu Tang, Qiuping Ding, Qiongbin Zhu, Lisan Zhang, Zhicai Chen, Hongfu Li, Wei Luo, Zhidong Cen, Jianzhong Sun, Minming Zhang, Jiawei Liang, Hongxi Zhang, Zhihan Yan, Yixin Emu, Xijing Zhang, Keyan Yu, Guanxun Cheng, Yadong Liu, Libo Zhang, Sven Haller, James Cole, Yuxin Li, Chao Wang, Peiyu Huang, Fang Xie, Hanzhang Lu, Dan Wu, Tengfei Guo, Xin Xu, Dengrong Jiang, Yaou Liu

**Affiliations:** Zhejiang Key Laboratory of Intelligent Sensing Technology and Advanced Medical Instrument and Key Laboratory for Biomedical Engineering of Ministry of Education, College of Biomedical Engineering & Instrument Science, Zhejiang University, Hangzhou, China; Department of Medical Imaging, Peking University Shenzhen Hospital, Shenzhen, China; Department of Radiology, Beijing Tiantan Hospital, Capital Medical University, Beijing, China; China National Clinical Research Center for Neurological Diseases, Beijing, China; Department of Radiology, The Second Affiliated Hospital, Zhejiang University School of Medicine, Hangzhou, China; Department of Psychiatry, School of Public Health, The Second Affiliated Hospital, Zhejiang University School of Medicine, Hangzhou, China; Department of Radiology, Huashan Hospital, Fudan University, Shanghai, China; The Second Affiliated Hospital and Yuying Children’s Hospital of Wenzhou Medical University, Wenzhou, China; Department of Nuclear Medicine & PET Center, Huashan Hospital, Fudan University, Shanghai, China; Institute of Neurological and Psychiatric Disorders, Shenzhen Bay Laboratory, Shenzhen, China; Department of Radiology, Johns Hopkins University School of Medicine, Baltimore, MD, USA; Department of Radiology, Shengjing Hospital of China Medical University, Shenyang, China; National Engineering Research Center of Advanced Magnetic Resonance Technologies for Diagnosis and Therapy, School of Biomedical Engineering, Shanghai Jiao Tong University, Shanghai, China; Department of Neurology, Sir Run Run Shaw Hospital, Zhejiang University School of Medicine, Hangzhou, China; Department of Neurology, The Second Affiliated Hospital, Zhejiang University School of Medicine, Hangzhou, China; Department of Radiology, Children’s Hospital, Zhejiang University School of Medicine, National Clinical Research Center for Child Health, Hangzhou, China; MR Scientific Collaboration, United Imaging Healthcare, Shanghai, China; Department of Radiology, Guiqian International General Hospital, Guiyang, China; Department of Radiology, Third Hospital of Heilongjiang, Beian, China; Centre d’Imagerie Médicale de Cornavin (CIMC), 1201 Geneva, Switzerland; Department of Computer Science, University College London, London, UK

**Author notes:** These authors co-correspond to this work. **Correspondence author:** Yaou Liu, M.D., Ph.D., Department of Radiology, Beijing Tiantan Hospital, Capital Medical University, Beijing, China. These authors contributed equally to this work (Zixuan Lin, Xiang Fan, Tianyu Gao, Shuyue Wang, Yiwen Hong).

## Abstract

Cerebral oxygen extraction fraction (OEF) reflects the balance between cerebral oxygen delivery and metabolic demand, but its normative evolution across the human lifespan remains unknown. Here we used rapid, non-contrast TRUST MRI to establish a multisite normative model of global cerebral OEF in 2,025 healthy individuals aged 0-93 years from 17 imaging sites. OEF increased from the neonatal period to middle adulthood, followed by a slower rise and plateau in later life, with the fastest change occurring during early development and no significant sex differences. Individual OEF deviation scores were associated with vascular risk burden in healthy adults. Applying the model to 885 patients revealed disease-related OEF alterations, including positive deviations in pediatric obstructive sleep apnea, autoimmune disorders, brain tumors, mild cognitive impairment and dementia. OEF deviation further tracked tumor grade and Ki-67 proliferation. These findings establish lifespan OEF charting as a scalable framework for individualized physiological neuroimaging.

## Introduction

The human brain constitutes 2% of the total body mass but consumes 20% of the oxygen^1^. Because the brain has minimal oxygen reserves, cerebral energy metabolism largely depends on the ability of the brain to extract oxygen from the incoming blood. Cerebral oxygen extraction fraction (OEF) is such a key physiological index that quantifies the fraction of oxygen extracted from arterial blood as it traverses the cerebral circulation^2^. Baseline cerebral OEF has been associated with neurovascular coupling and may serve as an important indicator of resting-state cerebral metabolic function. From a clinical perspective, OEF is uniquely informative because it reflects the regulated balance between oxygen delivery, vascular reserve and metabolic demand, and may reveal disease-related metabolic vulnerability before overt structural damage occurs. This physiological sensitivity makes OEF complementary to conventional structural MRI markers, such as regional volume loss or cortical atrophy, which primarily capture downstream anatomical consequences of disease. In addition, OEF provides direct hemodynamic and metabolic information that may become abnormal before detectable structural changes. Consistent with this role, altered OEF has been implicated in a wide range of neurological and systemic disorders, such as Alzheimer’s disease (AD)^3^, cerebral small vessel disease (cSVD)^4^, stroke^5^, and brain tumors^6^.

Despite its physiological and clinical relevance, the interpretation of cerebral OEF remains limited by the absence of a lifespan normative reference. OEF is shaped by development, aging, vascular status and disease, making it difficult to determine from an isolated measurement whether an individual falls within the expected physiological range or shows pathological deviation. Establishing normative OEF trajectories is therefore essential for transforming OEF from a group-level physiological measure into an individualized marker of brain metabolic vulnerability. Recently, normative modeling has emerged as a powerful strategy to address this type of problem in neuroimaging. For example, Bethlehem et al. constructed a brain chart of brain structural changes by aggregating the largest multisite structural MRI datasets, totaling 123,984 scans^7^. The application of this brain morphology charting in Chinese population was then reported^8^. Similarly, normative models were generated for white matter micro- and macrostructure, functional connectome and effective connectome^9–11^. Extending this approach to the field of cerebral physiological function, Zeng et al. established a perfusion chart to elucidate lifespan CBF dynamics^12^. Such approaches provide a quantitative reference for identifying atypical brain development, aging-related vulnerability and disease-associated alterations, and have increasingly been used to support individualized diagnosis, stratification and risk assessment^7, 8^.

However, to date, the potential of this normative modeling in OEF research remains unexplored, which could be largely attributed to the scarcity of non-invasive and reliable tools for OEF measurement. ^15^O-PET is currently the gold standard for measuring OEF but is rarely used due to the highly-invasive and complex procedure, making it unsuitable for large-scale multisite lifespan studies^13, 14^. Over the past decades, an active research topic in MRI field is to develop new techniques for noninvasive OEF measurement^15–30^. T_2_-relaxation-under-spin-tagging (TRUST) MRI is one of the most representative methods and showed a strong consistency (ICC = 0.90) with ^15^O-PET measurement in the same group of individuals^15, 31^. It does not require exogeneous tracer injection and provides a global measurement of OEF with a short scan time of 1.2min and an excellent reproducibility^32–35^, ready for large-scale lifespan and clinical studies.

Therefore, this study aimed to leverage this technical advance to develop a normative model for cerebral oxygen extraction. The lifespan trajectory of OEF from neonates to 93 years old was computed based on 2,025 TRUST datasets from 17 sites. We further assessed its clinical utility by examining OEF deviation z-scores in relation to vascular risk burden and disease-related alterations across 885 patients with pediatric and adult disorders. Finally, we demonstrated the stability of OEF normative model with a longitudinal test-retest study, a traveling study and a series of sensitivity analysis. As the first study to establish normative model for cerebral oxygen extraction, this work provides a robust framework for individualized assessments of OEF and has the potential to advance both neuroscience research and clinical applications.

## Results

### TRUST MRI datasets and OEF quantification

We first established a multisite TRUST MRI sampling resource for normative modeling of cerebral OEF. The reference cohort included 2,085 healthy control (HC) participants from 17 imaging sites, covering 16 sites in 8 cities in China and 1 site in the United States. A parallel clinical cohort included 978 patients from 9 sites within the same network, spanning pediatric and adult disorders. In addition, two prospective reliability datasets were included to assess longitudinal test-retest stability and inter-site reproducibility of TRUST-derived OEF. Together, this resource enabled us to construct lifespan OEF trajectories, quantify individual deviations from normative expectations, and examine disease-related alterations across heterogeneous clinical populations.

TRUST MRI quantifies global OEF by exploiting the strong dependence of venous blood T_2_ relaxation time on oxygenation level^36, 37^ (Figure 1a-b). Specifically, OEF can be derived using an established calibration model once the pure venous blood T_2_ is accurately measured^38^. TRUST MRI is a dedicated and carefully optimized sequence designed to isolate venous blood signals while minimizing contamination from surrounding tissue, thereby enabling robust estimation of venous oxygenation^15, 39^. Across all participating sites, acquisition parameters followed previously published protocols to ensure methodological consistency^33, 35^. Minor variations in echo time (TE) were present due to site-specific hardware limitations; however, these differences were small (< 2ms) and did not meaningfully affect signal quality or downstream OEF estimation.

**Figure 1.**
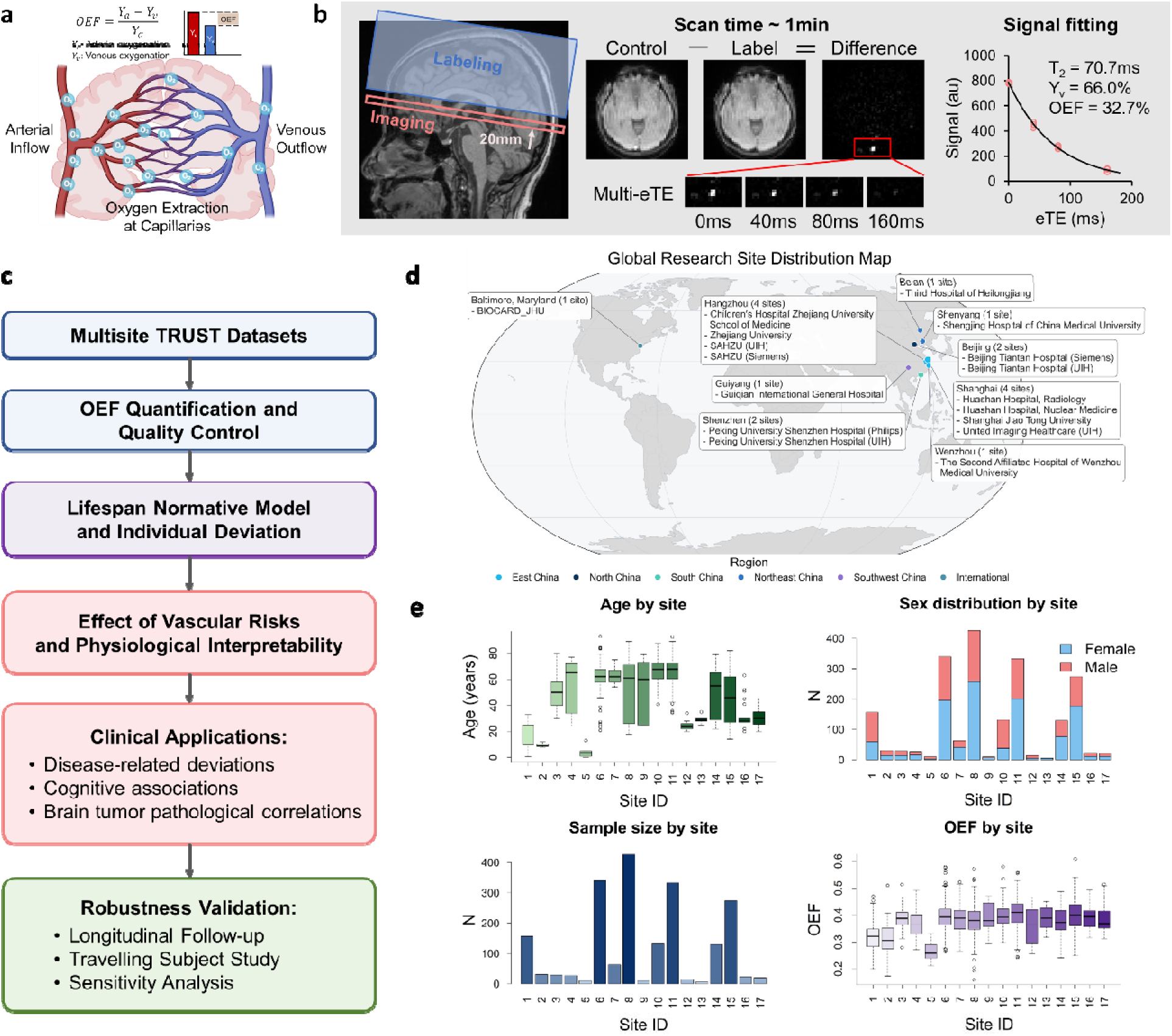
MRI of cerebral oxygen extraction fraction (OEF) and study design. (a) Definition of OEF. (b) TRUST MRI for rapid, non-invasive OEF quantification. (c) Study design and workflow. (d) Distribution map of 17 sites included in this study. (e) Distribution of age, sex, sample size and OEF across sites.

To further ensure data quality, an automatic quality control procedure was applied based on the 95% confidence interval of the measured blood T_2_ values (delta-R_2_). Scans with implausible T_2_ estimates, i.e. delta-R_2_ ≤ 10Hz were excluded. After quality control, the final dataset comprised 2,910 participants ranging aged 0 to 93 years, including 2,025 HCs and 885 patients. Detailed demographic information, including age distribution, sex, and site-specific characteristics, is summarized in Figure 1e.

### Normative trajectory of global cerebral oxygen extraction fraction across the lifespan

To characterize the lifespan trajectory of global cerebral OEF, we employed generalized additive models for location, scale, and shape (GAMLSS)^40^, which allow flexible modeling of age-related changes in both the mean and variability of OEF (Figure 2a-b and Supplementary Figure S1). Candidate models were evaluated across distribution families and spline complexities using the Bayesian information criterion (BIC). The optimal model used the Box-Cox t distribution (BCTo), with μ modeled as a B-spline function of age with 2 degrees of freedom, sex, and their interaction, with site included as a random effect; σ was modeled as a linear B-spline function of age with 1 degree of freedom and sex as a fixed effect, while ν and τ were specified as intercept-only terms. The results revealed a steady increase in global OEF from the neonatal period (OEF≈0.26) through middle adulthood (OEF≈0.36), followed by a markedly slower increase and eventual plateau in older age (OEF≈0.39). Inter-individual variability was quantified as the model-derived 5^th^-95^th^ percentile width at each age. This width changed by only 15% across adulthood (Supplementary Figure S3), supporting the notion of broadly stable inter-individual variability in global OEF during adulthood. No significant sex differences were observed across the lifespan.

**Figure 2.**
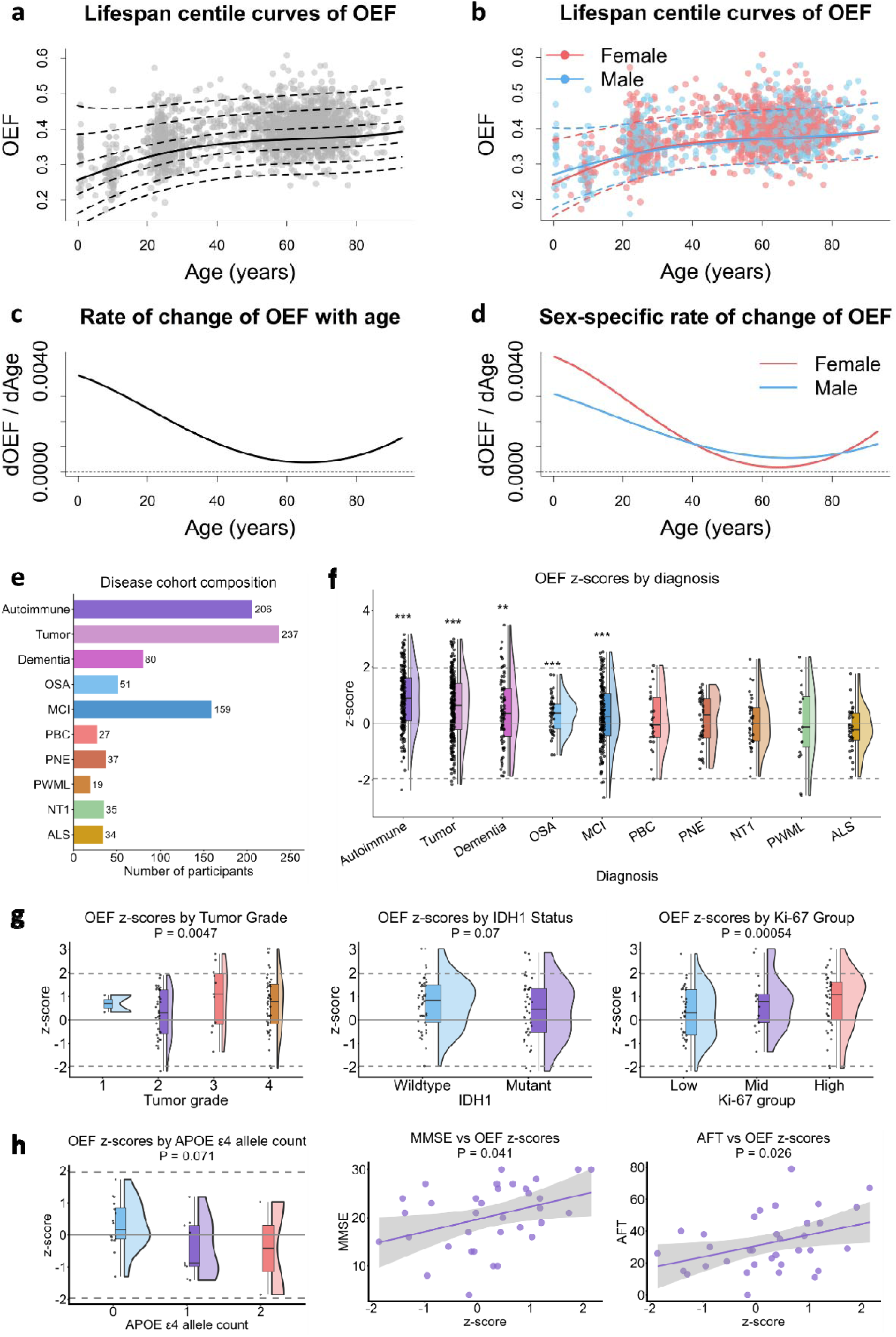
Evolution of OEF across the human lifespan and clinical application. (a) Lifespan centile curves of OEF. (b) Lifespan centile curves of OEF by sex. (c) Rate of change of OEF. (d) Sex-specific rate of change. (e) Composition of the disease cohort. (f) Deviation of OEF z-scores across different diseases. (g) Relationship between OEF z-scores and tumor grade, IDH1 mutation status and Ki-67 proliferation status in brain tumor patients. (h) Relationship between OEF z-scores and APOE genotype (number of ε4 alleles), MMSE score and AFT score.

Growth rate analyses (Figure 2c-d) provided further insight into the temporal dynamics of OEF. The rate of change was highest during early life (up to 0.004 per year), indicating a rapid increase in OEF during development. This increase progressively slowed with age, approaching near-zero levels at 60 years old, suggesting a plateau phase. In very late life, the growth rate exhibited a mild re-increase. Consistent with the mean trajectory, no sex differences were detected in growth rate patterns.

### Individual OEF deviation captures vascular risk burden in healthy adults

Having established the normative trajectory of OEF, we next asked whether individual OEF deviation scores capture biologically meaningful variation beyond normal aging. We first focused on vascular risk factors in neurologically normal participants because vascular comorbidities are known to influence cerebral hemodynamics^41–43^ and may represent an important source of subclinical OEF deviation before overt neurological disease. This analysis was therefore intended as a validation of the physiological interpretability of OEF z-scores and as a test of whether vascular risk burden contributes to higher-than-expected OEF in otherwise normal individuals.

OEF deviation z-scores were calculated relative to the normative model, providing a standardized measure of how much each individual deviated from the expected OEF value for their age. There were no significant differences in deviation scores across age, sex, or imaging sites for HC (Supplementary Figure S2), further supporting the robustness and generalizability of the normative model across demographic and technical factors.

Vascular risk information was available for 735 cognitively normal participants across 3 sites. A modified vascular risk score (VRS) was constructed by summing four commonly assessed vascular risk factors^44^: hypertension, hypercholesterolemia, diabetes, and overweight status defined as body mass index (BMI) > 25, adapted from prior composite vascular risk approaches. Each factor was coded as 0 (absent) or 1 (present, either recent or remote), yielding a total score ranging from 0 to 4, with higher scores indicating greater vascular risk burden. VRS was available in 552 participants with complete information. In addition, lifestyle-related variables, including smoking and alcohol consumption, were recorded for 470 participants, to explore potential associations between daily habits and cerebral oxygen extraction. The detailed distribution of individual risk factors and composite scores is presented in Supplementary Tables S1 and S2. A series of univariate linear regression analyses were conducted, with deviation score as the dependent variable and each risk factor or composite VRS as the independent variable.

A significant positive association between the modified VRS and individual OEF deviation scores was observed (β = 0.20, standard error (SE) = 0.038, P < 0.0001; Figure 3a), indicating that participants with a higher vascular risk burden tended to exhibit OEF above the normative trajectory. This association remained consistent when VRS was treated as a categorical variable, supporting a graded increase in OEF deviation with increasing vascular risk burden. For each vascular risk factor, the results showed that BMI status (P = 0.00034), hypertension (P = 0.00098), hyperlipidemia (P = 0.011), and diabetes (P = 0.0019) were all associated with increased individual deviation of OEF (Figure 3b). In the multivariable model including all four vascular risk components simultaneously (Supplementary Figure S4), BMI status showed the largest standardized association with OEF z-score (standardized β = 0.12, P = 0.0056), followed by hypertension (standardized β = 0.10, P = 0.023). Diabetes and hyperlipidemia showed positive but marginal associations with OEF z-score (diabetes: standardized β = 0.080, P = 0.059; hyperlipidemia: standardized β = 0.077, P = 0.072). However, smoking and alcohol consumption as well as lifestyle score showed no significant effect on OEF deviation (Figure 3c-d).

**Figure 3.**
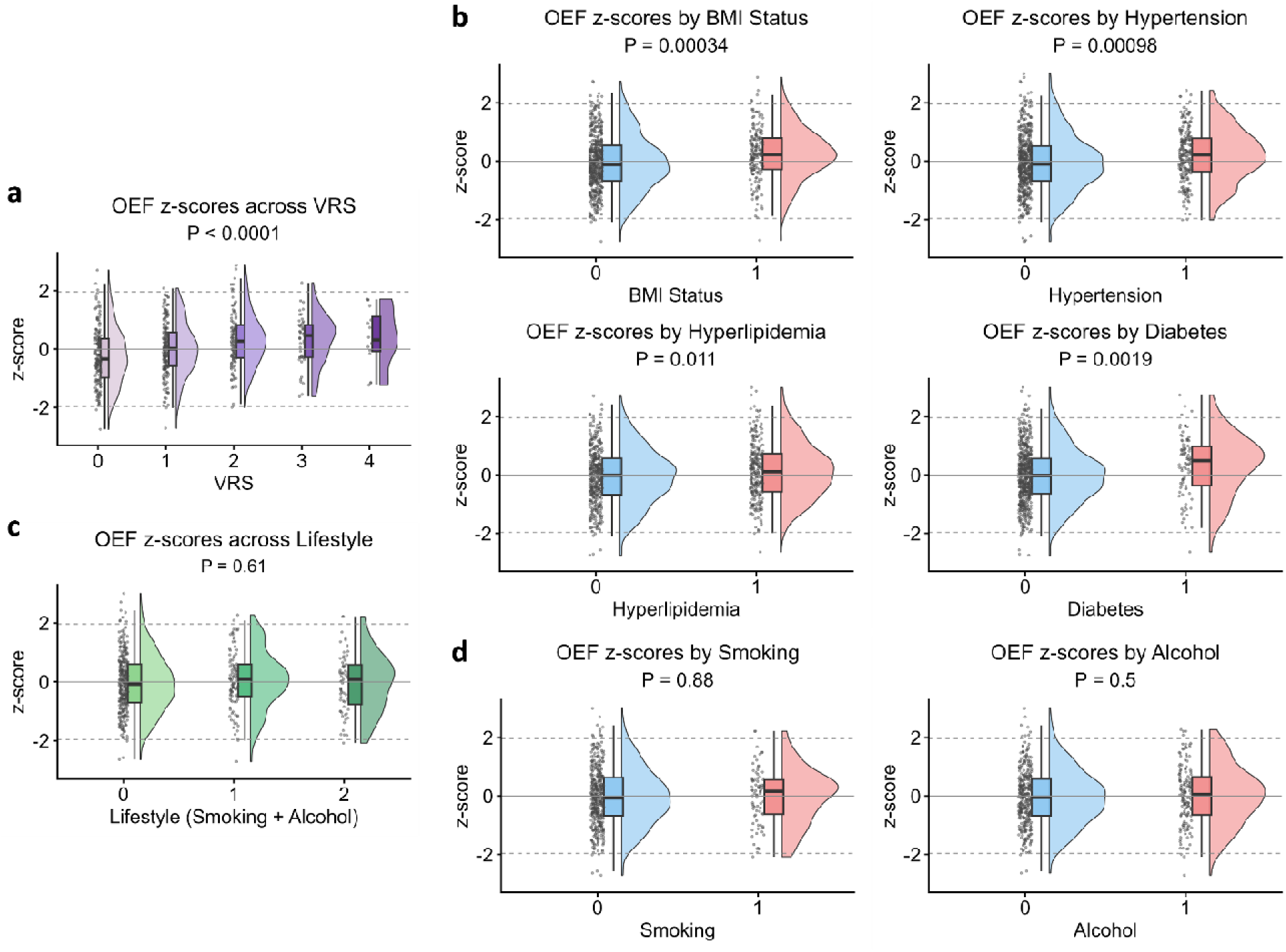
Effect of vascular risks and lifestyle on individual deviation of OEF. (a) Relationship between OEF z-scores and vascular risk score (VRS). (b) Relationship between OEF z-scores and individual vascular risk factors, including overweight status (BMI), hypertension, hyperlipidemia and diabetes. (c) Relationship between OEF z-scores and lifestyle score, constructed by the sum of smoking and alcohol consumption status. (d) Relationship between OEF z-scores and smoking or alcohol consumption status.

### Clinical relevance of the normative OEF framework

#### Task 1: Disease-specific pattern of OEF deviation

To evaluate the clinical utility of the current normative OEF model in a heterogeneous clinical context, we extended our analysis to include patient populations spanning multiple neurodevelopmental and neurological disorders. Patient data were collected from 8 imaging sites, all of which were included in the construction of the normative model, thereby ensuring methodological consistency and minimizing site-related confounds. As shown in Figure 2e, we included 3 pediatric disease categories, namely pediatric obstructive sleep apnea (OSA, N=51), primary nocturnal enuresis (PNE, N=37) and neonatal periventricular white matter lesions (PWML, N=19), allowing assessment of OEF alterations in neurodevelopmental and early-life brain disorders. In addition, we included 7 disease categories in adults, including mild cognitive impairment (MCI, N=159), dementia (N=80), autoimmune disease (N=206), brain tumor (N=237), narcolepsy-I (NT1, N=35), amyotrophic lateral sclerosis (ALS, N=34) and primary brain calcification (PBC, N=27). Together, these conditions encompass a broad spectrum of pathophysiological mechanisms, ranging from neurodevelopmental and early-life white matter abnormalities to sleep-related hypoxic stress, neuroinflammation, focal neoplastic processes and neurodegeneration. Detailed distribution of each disease population was listed in Supplementary Table S3. For each patient, individual OEF deviation scores were calculated as z-scores relative to the normative trajectory derived from healthy participants. Because a z-score of zero represents the expected OEF level for a healthy individual at the same age, we used one-sample t-tests against zero to determine whether each disease group showed a systematic deviation from the normative expectation (Figure 2f).

In pediatric populations, children with OSA showed a significant positive OEF deviation relative to the normative model (mean z = 0.31 ± 0.09 SE, Cohen’s d = 0.49, P = 0.00091, FDR q = 0.0023). Among adult neurological disorders, the strongest positive OEF deviations were observed in autoimmune disorders (mean z = 0.87 ± 0.07 SE, Cohen’s d = 0.84, P < 0.0001, FDR q < 0.0001) and brain tumors (mean z = 0.59 ± 0.07 SE, Cohen’s d = 0.52, P < 0.0001, FDR q < 0.0001), indicating substantially elevated cerebral oxygen extraction relative to normative expectations. Significant positive deviations were also identified in both MCI (mean z = 0.30 ± 0.08 SE, Cohen’s d = 0.29, P = 0.00038, FDR q = 0.0013) and dementia (mean z = 0.40 ± 0.14 SE, Cohen’s d = 0.33, P = 0.0047, FDR q = 0.0093), suggesting that elevated OEF is present across the cognitive impairment spectrum. By contrast, no significant OEF deviations were detected in PWML, PNE, PBC, NT1, or ALS.

#### Task 2: Linking OEF with tumor-related clinical phenotypes

To further investigate the clinical relevance of OEF deviation in brain tumors, we examined the association between individual OEF z-scores and key tumor-related pathological and clinical phenotypes in 90 patients, including tumor grade, Ki-67 proliferation index (in both continuous and categorical variables), isocitrate dehydrogenase 1 (IDH1) mutation status.

As illustrated in Figure 2g, linear regression analyses showed that OEF z-scores were significantly associated with tumor grade (β = 0.40, SE = 0.14, P = 0.0047) and Ki-67 level, both when Ki-67 was modeled as a continuous variable (β = 2.51, SE = 0.71, P = 0.00063) and as a categorical variable (β = 0.52, SE = 0.14, P = 0.00054). OEF z-scores also showed a trend-level association with IDH1 mutation status (β = −0.48, SE = 0.26, P = 0.070). That is, patients with higher tumor grade, higher Ki-67 proliferation and wildtype IDH1 status tended to exhibit elevated oxygen extraction fraction.

#### Task 3: Linking OEF with APOE genetics and cognitive performance

To further assess the relevance of OEF deviation to genetic and clinical features in neurodegenerative disease, we examined the association between individual OEF z-scores and APOE genotype and cognitive performance in 17 MCI and 16 dementia patients, including Mini-Mental State Examination (MMSE), Montreal Cognitive Assessment (MoCA), Animal Fluency Test (AFT), Symbol Digit Modalities Test (SDMT), and Neuropsychiatric Inventory (NPI).

As shown in Figure 2h, a univariate linear trend analysis treating APOE ε4 allele count as an ordinal predictor showed a marginal association with OEF deviation (β = −0.52, SE = 0.27, P = 0.071), with ε4 carriers tending to exhibit lower OEF z-scores. For the correlation with cognitive scores, after adjusting for vascular risk factors, lower OEF z-scores were significantly associated with worse MMSE (β = 2.34, SE = 1.09, P = 0.041) and AFT performance (β = 7.33, SE = 3.12, P = 0.026). In addition, trend-level associations were observed for SDMT (β = 5.46, SE = 2.82, P = 0.062) and NPI (β = −3.54, SE = 1.76, P = 0.054), whereas the association with MoCA was not significant (β = 1.54, SE = 1.07, P = 0.16).

### Stability of cerebral oxygen extraction fraction across time and space

As a multi-site study, rigorous evaluation of the temporal and spatial reproducibility of OEF measurement is essential to ensure the validity of the age-related trajectory and the robustness of multi-center normative modeling against longitudinal variability, physiological perturbations, and site-related factors such as scanner environment and operational differences. To this end, we prospectively designed two complementary validation datasets: one focusing on test-retest stability over a one-month interval (Figure 4a), and the other assessing inter-site reproducibility using a traveling-subject design (Figure 4b).

**Figure 4.**
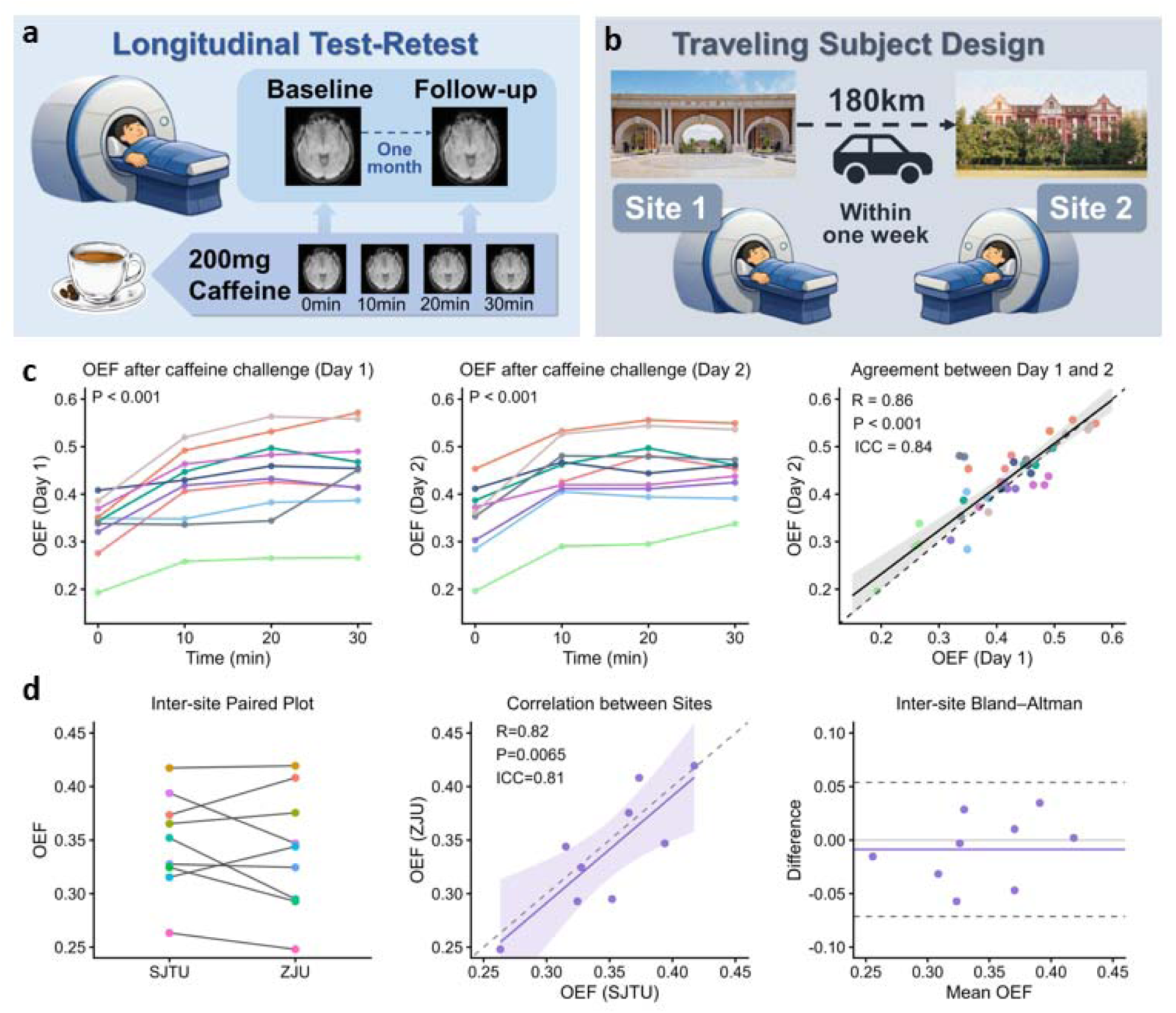
Stability of OEF measurement across space and time. (a) Longitudinal test-retest study design with caffeine challenge. (b) Travelling-subject study design. (c) OEF measurement in response to caffeine challenge on Day 1 and Day 2, as well as the agreement between two measurements. Each color represents one subject. (d) Agreement of OEF measurement between two sites.

To enhance the sensitivity of the stability assessment, 10 healthy participants underwent one baseline TRUST scan, followed by three additional scans after administration of 200 mg caffeine. Caffeine is a well-established vasoconstrictive agent and has been widely used as a physiological challenge in the technical validation of OEF-related MRI methods^45–47^. Inducing a predictable change in cerebral hemodynamics allowed us to test whether the technique could reliably detect both absolute OEF values and dynamic OEF changes. After one month, the same participants were scanned again using an identical protocol. As expected, OEF increased significantly after caffeine administration (P < 0.001 by linear-mixed effect model, Figure 4c). Importantly, the magnitude of OEF elevation was highly consistent between the baseline session and the one-month follow-up, indicating stable sensitivity to physiological modulation across time. Correlation analyses further demonstrated strong agreement between baseline and follow-up OEF measurements, with high correlation coefficients and excellent intraclass correlation coefficients (R = 0.86, ICC = 0.84, P < 0.001).

In addition to longitudinal stability, inter-site robustness was evaluated using a traveling-subject design. Another group of ten participants traveled between two imaging sites located in Hangzhou and Shanghai, namely ZJU and SJTU, approximately 180 kilometers apart, and underwent TRUST scans at both sites, with a mean inter-site scan interval of 24.2 ± 0.5 hours. This design enabled assessment of inter-site reproducibility. OEF measurements showed excellent agreement across the two sites (R = 0.82, ICC = 0.81, P = 0.0065, Figure 4d).

### Sensitivity analysis

The lifespan trajectory of cerebral OEF was further validated using a series of sensitivity analyses to assess the robustness of the normative patterns against potential confounding factors and analytical choices (see online Methods). Across all validation strategies, the estimated age-related OEF trajectories closely matched the main results (Extended Data Fig. 1). (i) To evaluate the potential influence of data quality, the analyses were repeated in a subset of participants meeting a stricter quality-control criterion for OEF estimation, i.e. delta-R_2_≤5Hz (N = 1976; Figure 5a). (ii) To mitigate the impact of uneven age distributions, a balanced resampling strategy was implemented to ensure uniform representation across age bins (N = 351 per resample, 500 resamples; Figure 5b). (iii) To assess reproducibility, a split-half analysis was performed by randomly dividing the cohort into two independent halves and estimating the normative OEF trajectory separately in each subset (Figure 5c). (iv) To examine sensitivity to sampling variability, bootstrap resampling was conducted with replacement (500 iterations; Figure 5d). (v) To evaluate the influence of individual imaging sites, a leave-one-site-out (LOSO) analysis was performed, in which the normative model was refit after sequentially excluding data from each site (Figure 5e).

**Figure 5.**
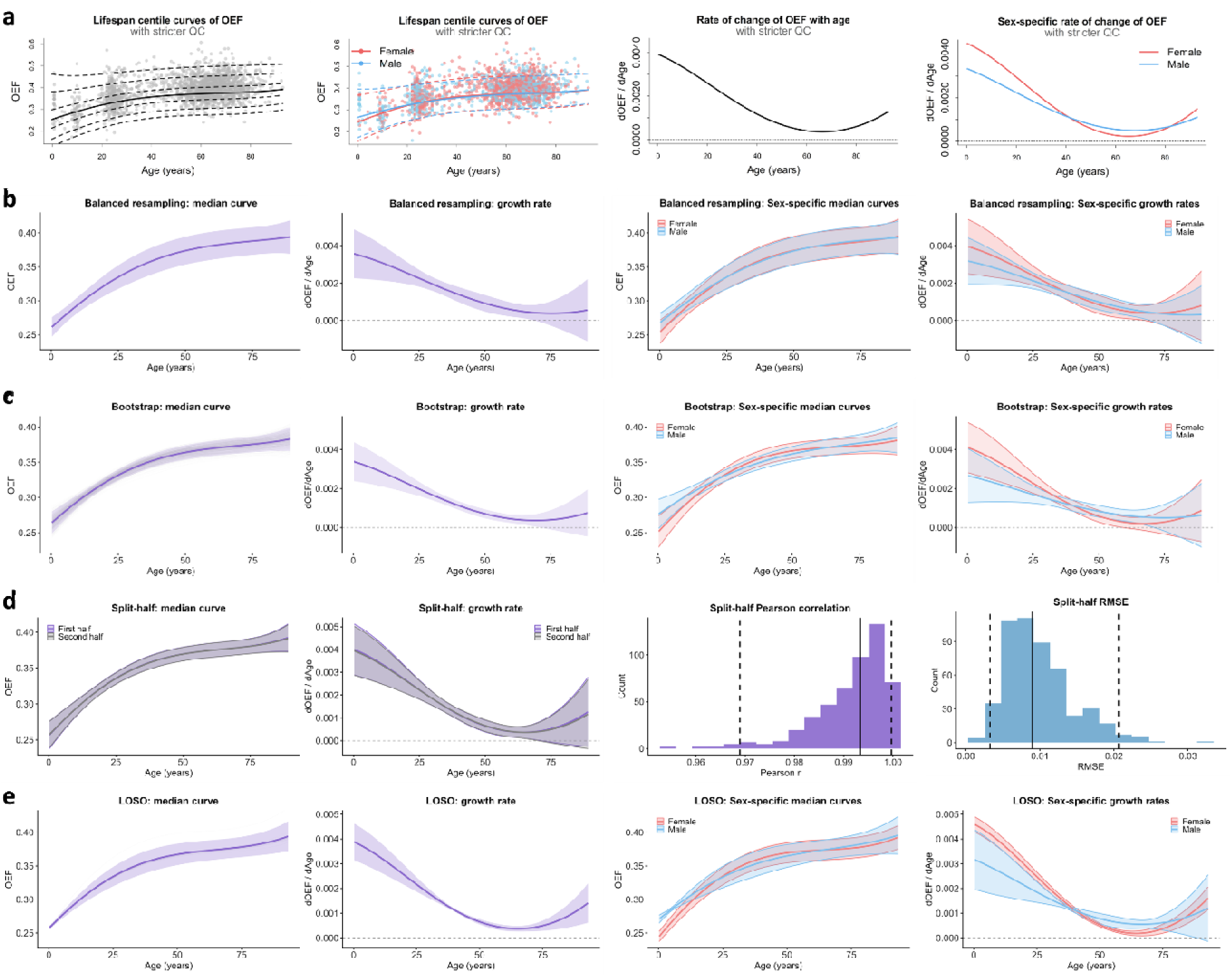
Sensitivity analysis for the normative OEF curves. (a) OEF centile curves and growth rates with stricter quality control (delta-R_2_≤5Hz). (b) OEF centile curves and growth rates with a balanced sampling design. (c) OEF centile curves and growth rates with bootstrap analysis. (d) OEF centile curves and growth rates with split-in-half analysis, along with the correlation of curves and RMSE between two halves. (e) OEF centile curves and growth rates with LOSO analysis.

The consistency between the OEF trajectories derived from the sensitivity analyses and the main model was also quantitatively assessed. Specifically, predicted median OEF values were sampled at one-year intervals across the lifespan, and Pearson’s correlation coefficients were calculated between the resulting curves (Supplementary Table 4). All sensitivity-derived trajectories exhibited high concordance with the main results (R ≈ 1.00, all P < 0.0001), indicating the robustness of the normative OEF trajectory to data quality, sampling strategy, and site composition. Similar levels of concordance were observed for the estimated rates of age-related change in OEF (R = 0.98-1.00, all P < 0.0001).

## Discussion

This study provides the first large, multisite characterization of cerebral oxygen extraction across the human lifespan. We mapped a nonlinear age-dependent OEF trajectory and established normative ranges of approximately 20-50%, enabling detection of individual deviations from expected values. Using rapid, non-contrast TRUST MRI, this framework can quantify global OEF within approximately one minute and is compatible with routine clinical MRI workflows due to its short scan time, minimal implementation burden and non-contrast nature. Supported by longitudinal test-retest, traveling-subject, and sensitivity analyses, TRUST-derived OEF provides a robust and clinically practical marker for characterizing normative brain physiology and disease-related deviations.

Previous evidence for age-related changes in OEF has been fragmented, with neonatal studies suggesting early increases^48^, scarce pediatric data^49^, and adult studies reporting age-related elevation in OEF^50–55^, limited by restricted age ranges, modest sample sizes and methodological heterogeneity. The present study extends this literature by delineating a continuous lifespan trajectory of OEF from the neonatal period to late adulthood in a large, unified multisite framework. We observed that global OEF increased steadily from the neonatal period through middle adulthood, followed by a slower increase and plateau in later life. The early rise likely supports the high metabolic demands of brain development, including synaptogenesis and myelination. As cerebral blood flow peaks around 7-10 years and declines thereafter^56^, the continued OEF increase may reflect greater reliance on oxygen extraction as cerebrovascular regulation, neurovascular coupling, and neural efficiency mature^57, 58^. In adulthood, higher OEF may partly reflect vascular aging^59^ and compensatory extraction in response to declining perfusion efficiency. consistent with its association with vascular risk burden^50, 51, 60–63^. The late-life plateau may represent a balance between vascular compensation and reduced metabolic demand^64, 65^, while the mild re-increase at very late age should be interpreted cautiously because of smaller sample size and potential survival bias.

Beyond defining normative OEF trajectories, our findings suggest that age- and sex-adjusted OEF deviation scores can identify disease-related metabolic and vascular abnormalities across pediatric and adult disorders, with elevated OEF potentially reflecting impaired oxygen delivery or compensatory extraction.

In pediatric OSA, positive OEF deviation was observed despite no significant T1-based structural differences, supporting the value of OEF as a complementary physiological marker beyond anatomical MRI. Given prior evidence of altered oxygen metabolism, reduced cerebrovascular reactivity, impaired autoregulation, and hypoperfusion in adult OSA^66–74^, elevated OEF may represent a compensatory response to reduced perfusion or insufficient flow adaptation during recurrent intermittent hypoxic stress.

Among adults, autoimmune disorders showed marked positive OEF deviation. This differs from prior multiple sclerosis (MS) studies reporting reduced OEF or CMRO_2_^75, 76^, likely reflecting differences in diagnosis and disease stage. Our cohort included heterogeneous autoimmune conditions which may have distinct metabolic profiles^77–79^, with many patients in early disease stages. Thus, elevated OEF may reflect active inflammation, reduced perfusion efficiency, or flow-metabolism mismatch, whereas reduced OEF may be more characteristic of chronic tissue injury or metabolic failure.

Brain tumors also exhibited marked positive OEF deviation, which was associated with tumor grade, Ki-67 proliferation, and IDH1 mutation, supporting a link between oxygen-metabolism-related imaging markers and malignant tumor biology^80–82^. However, because OEF reflects an integrated balance among oxygen delivery, vascular efficiency, metabolic demand, and tissue microenvironment, it should not be interpreted as tumor-specific. TRUST-derived OEF may instead serve as a complementary physiological marker alongside structural, perfusion, molecular, and pathological measures.

Within the cognitive impairment spectrum, both MCI and dementia showed elevated OEF. This finding contrasts with prior reports of reduced OEF in pure AD with limited vascular burden^3, 83^ and may instead reflect a compensatory response to coexisting hypoperfusion or vascular inefficiency, a pathophysiological context more representative of real-world cognitive impairment populations^84^, supported by recent studies in vascular-enriched and community-based aging cohorts that likewise reported elevated OEF in cognitive impairment^51, 60, 61^. After adjustment for vascular risk, OEF deviation was associated with poorer cognitive and neuropsychiatric performance, suggesting that OEF abnormalities may depend on the balance between vascular and AD-specific pathology^50^. APOE ε4 status was associated with lower OEF, in contrast to vascular risk, further supporting divergent physiological signatures^85^. Unlike T1-weighted MRI, which mainly reflects downstream atrophy, OEF may capture oxygen delivery-metabolism imbalance and help distinguish vascular-dominant from AD-dominant cognitive impairment.

Several limitations should be acknowledged. First, although this is the largest OEF dataset to date, its sample size remains modest compared with large-scale structural or functional MRI normative studies. Second, most imaging sites were located in China, and future studies should include more global centers to improve generalizability. Third, some clinical cohorts lacked detailed cognitive, functional, or longitudinal outcome measures, limiting direct assessment of clinical trajectories. Fourth, this study focused on OEF deviation alone and did not integrate deviations from other MRI features. Future work should test whether OEF provides complementary information to structural, perfusion, functional, or other physiological MRI deviation scores, and whether multimodal normative models improve prediction beyond univariate or non-normative MRI measures.

In conclusion, this study establishes a lifespan normative trajectory of cerebral oxygen extraction, providing a robust framework for quantifying individual variability and detecting physiological and pathological deviations. These findings support OEF-based models as sensitive tools for investigating brain metabolism and vascular health in research and clinical settings.

## Online Methods

### Participants and data collection

To delineate the normative trajectory of cerebral oxygen extraction in the human brain, we aggregated multisite neuroimaging datasets containing resting-state TRUST MRI scans. All recruitment procedures were approved by the local ethics committees, and all participants or their legal guardians provided written informed consent. A subset of participants received compensation for their time and effort, as detailed during the informed consent process. A total of 2,085 HCs and 978 individuals with various diseases participated in this study.

Healthy controls were identified using a harmonized cross-site definition rather than a single uniform recruitment pathway, because the normative cohort spanned a broad age range and was assembled from multiple site-specific studies. Participants were eligible for the healthy reference cohort if they had no known history of major neurological, psychiatric, cerebrovascular, or uncontrolled systemic disease, no contraindication to MRI, and no major structural brain abnormality on available anatomical MRI. Depending on the source cohort and age range, health status was determined by self-report, guardian report, review of medical history, clinical records, neuropsychological assessment, or study-specific clinical evaluation.

For neonatal participants (N = 24), eligibility was supported by neonatal clinical and imaging assessments. Neonates were included only if clinical MRI showed no evidence of brain injury or abnormality of the nervous system. In addition, birth-related laboratory examinations were required to be within the normal range, including umbilical cord blood gas analysis, liver function tests, routine blood examination, and serum ion analyses. Because the neonatal sample included both term-born and preterm-born infants, gestational age was recorded but was not used as a strict exclusion criterion.

For children beyond the neonatal period, adolescents, and younger-to-middle-aged adults aged ≤50 years recruited from community, school, university, or workplace-based sources (N = 660), healthy status was determined based on self-reported or guardian-reported medical history and anatomical MRI screening. Participants were included only if they reported no history of neurological disease or other major systemic disorders and showed no major structural abnormality on T1-weighted MRI.

For adults older than 50 years (N = 1401), healthy controls were recruited from three complementary sources. First, 351 participants were recruited directly from community-based cohorts and were included if they had no diagnosis of acute-stage stroke or transient ischemic symptoms, no diagnosis of dementia, no self-reported history of major neurological disease, major psychiatric disorder, or major systemic disorder, and no major structural abnormality on anatomical MRI. Second, 741 participants were selected from aging or dementia-oriented cohorts. For these participants, cognitively normal status was determined according to cohort-specific criteria, including MMSE- and CDR-based classification, or by clinical neurological evaluation confirming the absence of MCI, dementia, or other neurodegenerative diseases. Third, 309 participants were drawn from control groups of other disease-oriented case-control cohorts. These participants were included only if clinical evaluation confirmed the absence of the target disease and other major neurological, psychiatric, or systemic disorders, and if anatomical MRI showed no major structural abnormality. Across all older adult sources, age-related imaging findings commonly observed in older adults, such as mild-to-moderate white matter hyperintensities, were not used as absolute exclusion criteria unless they were judged locally to represent major structural pathology.

Exclusion criteria for the healthy reference cohort included diagnosed cognitive impairment or dementia, prior stroke, brain tumor, inflammatory or demyelinating neurological disease, major developmental brain abnormality, severe psychiatric illness, major uncontrolled systemic disease, or major structural brain abnormality on anatomical MRI. Thus, although recruitment strategies differed across sites and age ranges, all healthy controls met a shared minimum standard for neurological, cognitive, and structural normality.

After rigorous quality control, as described below, 2,025 HCs and 885 patients were included in the final statistical analysis. The patient cohort included 237 patients with brain tumors, 206 with autoimmune disease, 159 with mild cognitive impairment, 80 with dementia, 51 with obstructive sleep apnea, 37 with primary nocturnal enuresis, 35 with narcolepsy type 1, 34 with amyotrophic lateral sclerosis, 27 with primary brain calcification, and 19 with punctate white matter lesions. Age and sex information were collected for all participants, and the detailed distribution is listed in Supplementary Table S3.

### MRI experiments, image processing and quality control

#### (i) TRUST data collection

All MRI experiments were performed on 3T MRI systems. 760 HCs and 346 patients were scanned on Siemens Prisma or VIDA system; 874 HCs and 203 patients were scanned on Philips Achieva or Ingenia systems; 451 HCs and 429 patients were scanned on United Imaging 790, 880 or PMR systems. TRUST MRI was performed using the following parameters: TR = 3000 ms, TE = 3.6/3.9/5.1 ms for Siemens/Philips/United Imaging scanners, inversion time = 1022 ms, flip angle = 90°, FOV = 220 × 220 × 5 mm^3^, voxel size = 3.44 × 3.44 × 5 mm^3^, four effective TEs (1, 40, 80, and 160 ms) with a τCPMG of 10 ms, labeling thickness = 100 mm, and scan duration = 1.2/1.5/1.3 min for Siemens/Philips/United Imaging scanners.

#### (ii) TRUST image processing and quality control

TRUST imaging processing followed previous publications^33, 39, 50^. Briefly, a mono-exponential fitting of the TRUST MRI signal at the superior sagittal sinus as a function of eTE yielded blood T_2_. Blood T_2_ was in turn converted to venous oxygenation (Y_v_) using a calibration plot reported in Lu et al^38^. Then, the global OEF was calculated using the following equation:

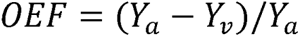

where Y_a_ is arterial oxygenation (assumed to be 98%).

The uncertainty of experimental measurement, represented by the 95% confidence interval of the estimated 1/T_2_ (delta-R_2_), was calculated for each participant. Datasets with delta-R_2_ greater than 10Hz were considered low reliability and were excluded from the analyses.

#### Modeling normative growth curves across the lifespan

To estimate normative growth patterns of OEF across the lifespan, we applied GAMLSS using the gamlss package (version 5.5.0) in R 4.5.1^40, 86^. A comprehensive model selection procedure was performed to jointly determine the optimal distribution family and model complexity (Supplementary Figure S1). Specifically, multiple candidate distribution families were evaluated, including NO, LO, BCCG, BCPE, BCTo, SHASH, and JSU, to account for potential skewness and kurtosis in the data. For each family, we systematically varied the degrees of freedom for the location (μ) and scale (σ) parameters, with μ modeled using B-splines of age (df = 2-6) and σ modeled with df = 1-4. All models included age and sex as fixed effects, with site modeled as a random effect in the location parameter. Model fitting was performed using a grid search across all combinations of distribution families and degrees of freedom, and only converged models were retained. The optimal model was selected as the one with low BIC as well as low degree of freedom for μ across all candidate models. Using the selected model, normative centile curves were estimated, and growth rates (dOEF/dAge) were derived via numerical differentiation of the predicted location parameter.

### Individual deviation scores and OEF patterns

#### (i) Calculation of individual deviation score

Individual deviation z-score was computed by transforming observed phenotype values through the cumulative distribution function of the fitted distribution, followed by conversion to the standard normal scale. For HCs, additional tenfold cross-validation was used to ensure that the individual predicted deviation scores were not included in the normative reference training. The normality of the z-score distributions was confirmed using Shapiro-Wilk test.

#### (ii) Association between individual deviation and vascular risks

Among normal participants, vascular risk factors were available in 735 individuals, including hypertension (N=730), hypercholesterolemia (N=634), diabetes (N=634), and overweight status (N=653). Smoking and alcohol information were available in 470 individuals. To investigate the relationship between vascular risk factors and deviations from normative trajectories, we conducted a series of univariate linear regression analyses using the calculated deviation score as the dependent variable, with each risk factor or the composite VRS as the independent variable.

### Clinical relevance of OEF-based normative models

#### (i) OEF deviation patterns in diseases

Disease-related alterations in cerebral oxygen extraction were also evaluated. Patient data were collected from 8 imaging sites included in the normative model to ensure methodological consistency and minimize site effects. For each subject, OEF deviation was quantified as a z-score relative to the normative trajectory estimated using GAMLSS. Then within each disease group, one-sample t-tests against zero (normative mean) were performed to assess deviations from normal. This normative-deviation approach preserves information from the full healthy reference cohort and provides a common reference scale across heterogeneous disease groups, avoiding the instability and reduced statistical power associated with repeatedly selecting smaller age- and sex-matched control subsets for each condition. Because multiple disease categories were tested, P values were adjusted across all ten disease-level comparisons using the Benjamini-Hochberg FDR procedure. Disease groups with q < 0.05 were considered to show significant deviation from the normative expectation.

#### (ii) Association between OEF deviation and clinical phenotypes in brain tumor and neurodegenerative diseases

To investigate the clinical relevance of OEF deviation, we examined the associations between individual OEF z-scores and disease-related phenotypes in both brain tumor and neurodegenerative cohorts using linear regression models, with OEF z-score as the dependent variable. In the brain tumor cohort, the variables of interest included tumor grade, Ki-67 proliferation index, and IDH1 mutation status. In the neurodegenerative cohort, the variables of interest included APOE genotype (coded for 0/1/2 ε4 alleles) and a range of cognitive, neuropsychiatric and functional measures. Continuous variables were modeled as continuous predictors and categorical variables as factor predictors. Because the brain tumor cohort was more heterogeneous and had a relatively larger sample size (N=89), these models were adjusted for age, sex, and VRS. In contrast, given the smaller sample size (N=33) and relatively narrower age distribution of the neurodegenerative cohort, and because OEF z-scores had already been standardized for age and sex using the GAMLSS normative model, neurodegenerative analyses were adjusted for VRS only to account for residual vascular confounding while avoiding overfitting.

### Stability analysis of OEF measurement across space and time

#### (i) Longitudinal OEF with one-month follow-up

To assess the longitudinal consistency and sensitivity to physiological modulation of TRUST-derived OEF measurements, we conducted a test-retest study with caffeine challenge in ten healthy participants. Each participant underwent an identical scanning protocol at two time points separated by one month. During each session, participants first received a baseline TRUST scan followed by three additional scans (10, 20 and 30 min) after oral administration of 200 mg caffeine, a well-characterized vasoconstrictive agent that reliably modulates cerebral hemodynamics. This within-session pharmacological challenge allowed evaluation of the technique’s sensitivity to detect dynamic physiological changes, while the one-month interval between sessions permitted assessment of longitudinal measurement consistency. Statistical analyses included paired comparisons of OEF values before and after caffeine administration, correlation analyses between baseline and follow-up measurements, and calculation of intraclass correlation coefficients to quantify measurement agreement across time points.

#### (ii) Inter-site reproducibility of OEF with traveling subject

To evaluate the inter-site reproducibility of TRUST-derived OEF measurements, we employed a traveling-subject design. Ten healthy participants traveled between two imaging sites located in Shanghai and Hangzhou (namely SJTU and ZJU, approximately 180 kilometers apart). All scans were completed within one week, and each participant’s scans were scheduled at the same time of day across sites and sessions to control for potential diurnal effects on OEF. At each site, participants underwent identical TRUST scanning protocols. Statistical analyses included correlation analyses and intraclass correlation coefficient calculations to quantify measurement agreement across sessions within the same site and across different sites.

### Sensitivity analysis of normative models

To further validate the lifespan normative trajectory of OEF, we conducted a series of sensitivity analyses to assess the robustness of the model with respect to data quality, age imbalance, site-related variability, and model reproducibility.

#### (i) Stricter quality control for OEF data

To ensure the robustness of our findings against variations in OEF data quality, we repeated the analysis in a subset of participants meeting a stricter quality-control criterion for OEF estimation, defined as delta-R_2_ ≤ 5 Hz. A normative model was re-fitted in this subset using the same modeling framework as in the main analysis, and the resulting trajectory was compared with that derived from the full cohort.

#### (ii) Balanced resampling analysis

To reduce the influence of uneven age distribution across the lifespan, we performed a balanced resampling analysis for global OEF. The cohort was divided into consecutive 10-year age bins spanning 0 to 90 years. In each iteration, random subsamples were drawn such that each age bin contributed the same number of participants, equal to the size of the smallest age bin. This procedure was repeated 500 times. Within each iteration, a GAMLSS model including age, sex, and a random site effect was re-fitted, and sex-averaged centile curves were derived using population-level prediction. The mean trajectory across resamples and its 95% confidence interval, defined as mean ± 1.96 SD, were then calculated for both the median trajectory and its first derivative. In addition, the agreement between each resampled median curve and the main model was quantified by correlation analysis.

#### (iii) Split-half analysis

To assess model reproducibility, we performed a split-half analysis by randomly dividing the full dataset into two halves within each site, thereby preserving site representation. For each half, the same fixed GAMLSS model structure was fitted independently, including age, sex, and a random site effect. Median centile curves and their first derivatives were derived for both halves using population-level prediction. Agreement between the two halves was quantified by Pearson correlation and root-mean-square error (RMSE) of the median trajectories. The entire split-half procedure was repeated 500 times, and summary statistics of these agreement metrics were obtained across repetitions.

#### (iv) Bootstrap resampling analysis

To assess robustness to sampling variability, we performed a stratified bootstrap analysis with 500 repetitions. Resampling was conducted with replacement within strata defined by 10 equally spaced age intervals and sex, thereby preserving the age-sex structure of the original cohort. For each bootstrap sample, the GAMLSS model was re-fitted, and the sex-averaged median trajectory was estimated. From the distribution of 500 bootstrap-derived trajectories, we calculated 95% confidence intervals for both the median trajectory and its first derivative. The ages corresponding to the peak and trough of the growth-rate curve were also extracted from each bootstrap iteration to summarize variability in age-related change patterns.

#### (v) Leave-one-site-out (LOSO) analysis

To evaluate the influence of individual imaging sites, we performed a leave-one-site-out (LOSO) analysis. In each iteration, data from one site were excluded, and the same GAMLSS model was re-fitted using the remaining data. Sex-averaged median trajectories and their first derivatives were then estimated using population-level prediction. This process was repeated across all sites. The mean and standard deviation of the resulting trajectories were used to derive 95% confidence intervals for both the median trajectory and growth rate.

## Data availability

Data used in this study are from private cohort studies, therefore we have no permission at current stage to distribute the data for public use. However, many datasets are available upon request to the corresponding author Dr. You Liu.

## Code availability

All code used for the statistical analyses and figure generation in this study is publicly available at GitHub (https://github.com/zixuanlin14zju/TRUST_Lifespan).

## Acknowledgements

We acknowledge all the contributors who provided magnetic resonance and clinical data. The work was supported by the Key Program of the National Natural Science Foundation of China (82330057), National Science Foundation of China (82302144, 32427802, 62401363), the Beijing Outstanding Young Scientist Program (JWZQ20240101025), Beijing Hospital Management Center-Climb Plan (DFL20220503), Beijing Young Scholars (NO.026), Shanghai Pujiang Program (24PJA047) and Xiaomi Young Scholar Program.

## Author Contribution

Study Design: Z. Lin, T. Gao, Y. Hong, H. Lu, D. Jiang, Yaou. Liu.

Data collection: Z. Lin, X. Fan, T. Gao, S. Wang, Y. Hong, Y. Yan, J. Liu, Y. Fu, T. Hua, Y. Cai, G. Lu, Y. Qi, B. Yu, Z. Zhuo, J. Wu, D. Ge, Q. Xu, Y. Hu, C. Xiong, W. Liu, R. Tang, Q. Ding, Q. Zhu, L. Zhang, Z. Chen, H. Li, W. Luo, Z. Cen, J. Sun, M. Zhang, J. Liang, H. Zhang, Z. Yan, Y. Emu, X. Zhang, K. Yu, G. Cheng, Yadong. Liu, L. Zhang, D. Wu, Y. Li, C. Wang, P. Huang, F. Xie, H. Lu, T. Guo, X. Xu, D. Jiang, and Yaou. Liu

Data analysis: Z. Lin, X. Fan, T. Gao, S. Wang, Y. Hong, Y. Yan, J. Liu, G. Lu, Y. Qi, B. Yu, J. Wu, D. Ge, Q. Xu, Y. Hu, C. Xiong, W. Liu, D. Jiang, and Y. Liu

Statistical analysis: Z. Lin, Y. Hong, D. Ge

Paper writing: Z. Lin and D. Jiang

Paper editing: Z. Lin, Z. Zhuo, S. Haller, J. Cole, H. Lu, Yaou. Liu

## References

1. Rolfe, D.F. & Brown, G.C. Cellular energy utilization and molecular origin of standard metabolic rate in mammals. Physiol Rev 77, 731–758 (1997).

2. Jiang, D. & Lu, H. Cerebral oxygen extraction fraction MRI: Techniques and applications. Magn Reson Med 88, 575–600 (2022).

3. Thomas, B.P., et al. Reduced global brain metabolism but maintained vascular function in amnestic mild cognitive impairment. J Cereb Blood Flow Metab 37, 1508–1516 (2017).

4. Zhang, R., et al. Oxygen extraction fraction in small vessel disease: relationship to disease burden and progression. Brain 148, 1950–1962 (2025).

5. Fan, A.P., et al. Elevated brain oxygen extraction fraction measured by MRI susceptibility relates to perfusion status in acute ischemic stroke. J Cereb Blood Flow Metab 40, 539–551 (2020).

6. Stadlbauer, A., et al. MR Imaging-derived Oxygen Metabolism and Neovascularization Characterization for Grading and IDH Gene Mutation Detection of Gliomas. Radiology 283, 799–809 (2017).

7. Bethlehem, R.A.I., et al. Brain charts for the human lifespan. Nature 604, 525–533 (2022).

8. Zhuo, Z., et al. Charting brain morphology in international healthy and neurological populations. Nature Neuroscience 29, 420–434 (2026).

9. Sun, L., et al. Human lifespan changes in the brain’s functional connectome. Nature Neuroscience 28, 891–901 (2025).

10. Li, G., et al. The effective connectome over a century of human life. Communications Biology 8, 1638 (2025).

11. Kim, M.E., et al. White matter micro- and macrostructure brain charts for the human lifespan. Nature (2026).

12. Zeng, X., et al. Normative Cerebral Perfusion Across the Lifespan. ArXiv (2025).

13. Mintun, M.A., Raichle, M.E., Martin, W.R. & Herscovitch, P. Brain oxygen utilization measured with O-15 radiotracers and positron emission tomography. J Nucl Med 25, 177–187 (1984).

14. Fan, A.P., et al. Quantification of brain oxygen extraction and metabolism with [(15)O]-gas PET: A technical review in the era of PET/MRI. Neuroimage 220, 117136 (2020).

15. Lu, H. & Ge, Y. Quantitative evaluation of oxygenation in venous vessels using T2-Relaxation-Under-Spin-Tagging MRI. Magn Reson Med 60, 357–363 (2008).

16. He, X. & Yablonskiy, D.A. Quantitative BOLD: mapping of human cerebral deoxygenated blood volume and oxygen extraction fraction: default state. Magnetic Resonance in Medicine: An Official Journal of the International Society for Magnetic Resonance in Medicine 57, 115–126 (2007).

17. Wehrli, F.W., Fan, A.P., Rodgers, Z.B., Englund, E.K. & Langham, M.C. Susceptibility-based time-resolved whole-organ and regional tissue oximetry. NMR in Biomedicine 30, e3495 (2017).

18. Fan, A.P., et al. Quantitative oxygenation venography from MRI phase. Magnetic Resonance in Medicine 72, 149–159 (2014).

19. Jiang, D., et al. Vessel-specific quantification of neonatal cerebral venous oxygenation. Magn Reson Med 82, 1129–1139 (2019).

20. Bolar, D.S., Rosen, B.R., Sorensen, A.G. & Adalsteinsson, E. QUantitative Imaging of eXtraction of oxygen and TIssue consumption (QUIXOTIC) using venular-targeted velocity-selective spin labeling. Magn Reson Med 66, 1550–1562 (2011).

21. Guo, J. & Wong, E.C. Venous oxygenation mapping using velocity-selective excitation and arterial nulling. Magn Reson Med 68, 1458–1471 (2012).

22. Jain, V., Langham, M.C. & Wehrli, F.W. MRI estimation of global brain oxygen consumption rate. J Cereb Blood Flow Metab 30, 1598–1607 (2010).

23. Haacke, E.M., Tang, J., Neelavalli, J. & Cheng, Y.C. Susceptibility mapping as a means to visualize veins and quantify oxygen saturation. J Magn Reson Imaging 32, 663–676 (2010).

24. Zhang, J., et al. Quantitative mapping of cerebral metabolic rate of oxygen (CMRO2) using quantitative susceptibility mapping (QSM). Magn Reson Med 74, 945–952 (2015).

25. Cho, J., et al. Cerebral metabolic rate of oxygen (CMRO(2)) mapping by combining quantitative susceptibility mapping (QSM) and quantitative blood oxygenation level-dependent imaging (qBOLD). Magn Reson Med 80, 1595–1604 (2018).

26. An, H. & Lin, W. Impact of intravascular signal on quantitative measures of cerebral oxygen extraction and blood volume under normo- and hypercapnic conditions using an asymmetric spin echo approach. Magn Reson Med 50, 708–716 (2003).

27. He, X. & Yablonskiy, D.A. Quantitative BOLD: mapping of human cerebral deoxygenated blood volume and oxygen extraction fraction: default state. Magn Reson Med 57, 115–126 (2007).

28. Gauthier, C.J. & Hoge, R.D. Magnetic resonance imaging of resting OEF and CMRO_ using a generalized calibration model for hypercapnia and hyperoxia. Neuroimage 60, 1212–1225 (2012).

29. Bulte, D.P., et al. Quantitative measurement of cerebral physiology using respiratory-calibrated MRI. Neuroimage 60, 582–591 (2012).

30. Wise, R.G., Harris, A.D., Stone, A.J. & Murphy, K. Measurement of OEF and absolute CMRO2: MRI-based methods using interleaved and combined hypercapnia and hyperoxia. Neuroimage 83, 135–147 (2013).

31. Jiang, D., et al. Validation of T(2) -based oxygen extraction fraction measurement with (15) O positron emission tomography. Magn Reson Med 85, 290–297 (2021).

32. Liu, P., Xu, F. & Lu, H. Test-retest reproducibility of a rapid method to measure brain oxygen metabolism. Magn Reson Med 69, 675–681 (2013).

33. Jiang, D., et al. Cross-vendor harmonization of T(2) -relaxation-under-spin-tagging (TRUST) MRI for the assessment of cerebral venous oxygenation. Magn Reson Med 80, 1125–1131 (2018).

34. Liu, P., et al. Multisite evaluations of a T2 -relaxation-under-spin-tagging (TRUST) MRI technique to measure brain oxygenation. Magn Reson Med 75, 680–687 (2016).

35. Eldirdiri, A., et al. Toward vendor-independent measurement of cerebral venous oxygenation: Comparison of TRUST MRI across three major MRI manufacturers and association with end-tidal CO(2). NMR Biomed 36, e4990 (2023).

36. van Zijl, P.C., et al. Quantitative assessment of blood flow, blood volume and blood oxygenation effects in functional magnetic resonance imaging. Nat Med 4, 159–167 (1998).

37. Li, W. & van Zijl, P.C.M. Quantitative theory for the transverse relaxation time of blood water. NMR Biomed 33, e4207 (2020).

38. Lu, H., et al. Calibration and validation of TRUST MRI for the estimation of cerebral blood oxygenation. Magn Reson Med 67, 42–49 (2012).

39. Xu, F., Uh, J., Liu, P. & Lu, H. On improving the speed and reliability of T2-relaxation-under-spin-tagging (TRUST) MRI. Magn Reson Med 68, 198–204 (2012).

40. Borghi, E., et al. Construction of the World Health Organization child growth standards: selection of methods for attained growth curves. Statistics in Medicine 25, 247–265 (2006).

41. Uchino, K., et al. Increased cerebral oxygen metabolism and ischemic stress in subjects with metabolic syndrome-associated risk factors: preliminary observations. Transl Stroke Res 1, 178–183 (2010).

42. de la Torre, J.C. Cerebral hemodynamics and vascular risk factors: setting the stage for Alzheimer’s disease. J Alzheimers Dis 32, 553–567 (2012).

43. Ma, Y., et al. Changes in Cerebral Hemodynamics and Progression of Subclinical Vascular Brain Disease: A Population-Based Cohort Study. Stroke 56, 95–104 (2025).

44. Gottesman, R.F., et al. Association Between Midlife Vascular Risk Factors and Estimated Brain Amyloid Deposition. Jama 317, 1443–1450 (2017).

45. Xu, F., Liu, P., Pekar, J.J. & Lu, H. Does acute caffeine ingestion alter brain metabolism in young adults? Neuroimage 110, 39–47 (2015).

46. Lin, Z., et al. Vessel-specific quantification of cerebral venous oxygenation with velocity-encoding preparation and rapid acquisition. Magn Reson Med 92, 782–791 (2024).

47. Hong, Y., et al. Accelerated 3D Mapping of Whole Brain Venous Oxygenation With Nested-Shift Under Sampling and ESPIRIT Reconstruction. Magn Reson Med 95, 2023–2034 (2026).

48. Lin, Z., Wu, D., Jiang, D., Lu, H. & Qi, Y. Altered cerebral oxygen extraction and metabolism in preterm neonates and the relationship to anemia: A noncontrast MRI study. Annals of the Child Neurology Society 2, 269–280 (2024).

49. Takahashi, T., Shirane, R., Sato, S. & Yoshimoto, T. Developmental changes of cerebral blood flow and oxygen metabolism in children. AJNR Am J Neuroradiol 20, 917–922 (1999).

50. Jiang, D., et al. Brain Oxygen Extraction Is Differentially Altered by Alzheimer’s and Vascular Diseases. J Magn Reson Imaging 52, 1829–1837 (2020).

51. Lin, Z., et al. Longitudinal changes in brain oxygen extraction fraction (OEF) in older adults: Relationship to markers of vascular and Alzheimer’s pathology. Alzheimers Dement 19, 569–577 (2023).

52. Peng, S.L., et al. Age-related increase of resting metabolic rate in the human brain. Neuroimage 98, 176–183 (2014).

53. Jiang, D., et al. Normal variations in brain oxygen extraction fraction are partly attributed to differences in end-tidal CO(2). J Cereb Blood Flow Metab 40, 1492–1500 (2020).

54. Lu, H., et al. Alterations in cerebral metabolic rate and blood supply across the adult lifespan. Cereb Cortex 21, 1426–1434 (2011).

55. Catchlove, S.J., et al. An investigation of cerebral oxygen utilization, blood flow and cognition in healthy aging. PLoS One 13, e0197055 (2018).

56. Zhao, M.Y., et al. Measuring Quantitative Cerebral Blood Flow in Healthy Children: A Systematic Review of Neuroimaging Techniques. J Magn Reson Imaging 59, 70–81 (2024).

57. Baller, E.B., et al. Developmental coupling of cerebral blood flow and fMRI fluctuations in youth. Cell Rep 38, 110576 (2022).

58. Tau, G.Z. & Peterson, B.S. Normal development of brain circuits. Neuropsychopharmacology 35, 147–168 (2010).

59. Ding, Y., et al. Comprehensive human proteome profiles across a 50-year lifespan reveal aging trajectories and signatures. Cell 188, 5763–5784.e5726 (2025).

60. Song, J., et al. Elevated brain oxygen extraction fraction (OEF) as a potential marker for vascular cognitive impairment and dementia. Alzheimer’s & Dementia 21, e109994 (2025).

61. Yan, Y., et al. Cerebral oxygen extraction and blood flow in community-based older adults: associations with white matter hyperintensity and neurocognitive function. Brain Communications 8, fcag056 (2026).

62. King, K.S., et al. Detrimental effect of systemic vascular risk factors on brain hemodynamic function assessed with MRI. Neuroradiol J 31, 253–261 (2018).

63. Lee, D., Choi, Y., Jeong, E., Eun Park, J. & Kim, H.S. Relationship between cerebrovascular risk factors and oxygen metabolic stress in a cognitively impaired population: Dynamic susceptibility contrast-derived oxygen parametric analysis. J Cereb Blood Flow Metab 45, 2360–2369 (2025).

64. Walker, J.M., et al. The Spectrum of Alzheimer-Type Pathology in Cognitively Normal Individuals. J Alzheimers Dis 91, 683–695 (2023).

65. Alafuzoff, I. & Libard, S. Ageing-Related Neurodegeneration and Cognitive Decline. Int J Mol Sci 25 (2024).

66. Rodgers, Z.B., et al. Cerebral metabolic rate of oxygen in obstructive sleep apnea at rest and in response to breath-hold challenge. J Cereb Blood Flow Metab 36, 755–767 (2016).

67. Wu, P.H., et al. MRI evaluation of cerebral metabolic rate of oxygen (CMRO(2)) in obstructive sleep apnea. J Cereb Blood Flow Metab 42, 1049–1060 (2022).

68. Urbano, F., Roux, F., Schindler, J. & Mohsenin, V. Impaired cerebral autoregulation in obstructive sleep apnea. Journal of Applied Physiology 105, 1852–1857 (2008).

69. Morgan, B.J., et al. Effects of sleep-disordered breathing on cerebrovascular regulation: A population-based study. Am J Respir Crit Care Med 182, 1445–1452 (2010).

70. Jensen, M.L.F., Vestergaard, M.B., Tønnesen, P., Larsson, H.B.W. & Jennum, P.J. Cerebral blood flow, oxygen metabolism, and lactate during hypoxia in patients with obstructive sleep apnea. Sleep 41 (2018).

71. Yan, L., et al. Altered regional cerebral blood flow in obstructive sleep apnea is associated with sleep fragmentation and oxygen desaturation. J Cereb Blood Flow Metab 41, 2712–2724 (2021).

72. Nie, S., et al. Resting cerebral blood flow alteration in severe obstructive sleep apnoea: an arterial spin labelling perfusion fMRI study. Sleep Breath 21, 487–495 (2017).

73. Li, X., et al. Altered cerebral blood flow and white matter during wakeful rest in patients with obstructive sleep apnea: a population-based retrospective study. Br J Radiol 96, 20220867 (2023).

74. Innes, C.R., Kelly, P.T., Hlavac, M., Melzer, T.R. & Jones, R.D. Decreased Regional Cerebral Perfusion in Moderate-Severe Obstructive Sleep Apnoea during Wakefulness. Sleep 38, 699–706 (2015).

75. Ge, Y., et al. Characterizing brain oxygen metabolism in patients with multiple sclerosis with T2-relaxation-under-spin-tagging MRI. J Cereb Blood Flow Metab 32, 403–412 (2012).

76. Fan, A.P., et al. Quantitative oxygen extraction fraction from 7-Tesla MRI phase: reproducibility and application in multiple sclerosis. J Cereb Blood Flow Metab 35, 131–139 (2015).

77. Bai, S., et al. Multi-tracer PET and MR imaging visualize distinct metabolic and inflammatory profiles in the white matter of NMOSD and MOGAD. Neurotherapeutics 22, e00720 (2025).

78. Rashid, W., et al. Abnormalities of cerebral perfusion in multiple sclerosis. J Neurol Neurosurg Psychiatry 75, 1288–1293 (2004).

79. Liu, X., et al. The Clinical Value of 18F-FDG-PET in Autoimmune Encephalitis Associated With LGI1 Antibody. Frontiers in Neurology Volume 11 -2020 (2020).

80. Tóth, V., et al. MR-based hypoxia measures in human glioma. J Neurooncol 115, 197–207 (2013).

81. Saitta, L., et al. Signal intensity in T2’ magnetic resonance imaging is related to brain glioma grade. Eur Radiol 21, 1068–1076 (2011).

82. Oughourlian, T.C., et al. Relative oxygen extraction fraction (rOEF) MR imaging reveals higher hypoxia in human epidermal growth factor receptor (EGFR) amplified compared with non-amplified gliomas. Neuroradiology 63, 857–868 (2021).

83. Lajoie, I., et al. Application of calibrated fMRI in Alzheimer’s disease. Neuroimage Clin 15, 348–358 (2017).

84. Takeda, K., Noiri, A., Nakajima, T., Kobayashi, T. & Tarucha, S. Quantum error correction with silicon spin qubits. Nature 608, 682–686 (2022).

85. Lin, Z., et al. Brain Oxygen Extraction by Using MRI in Older Individuals: Relationship to Apolipoprotein E Genotype and Amyloid Burden. Radiology 292, 140–148 (2019).

86. Rigby, R.A. & Stasinopoulos, D.M. Generalized Additive Models for Location, Scale and Shape. Journal of the Royal Statistical Society Series C: Applied Statistics 54, 507–554 (2005).

