## Supplementary Figure S1 for "Oxygen extraction fraction brain charts for human lifespan and application for brain disorders"

Supplementary Table 1. Availability and completeness of vascular risk factor information

| **Variable** | **N** | **%** |
| --- | --- | --- |
| Vascular risk data available for ≥1 VRS component | 735 | 39.2 |
| Complete data for all 4 VRS components | 552 | 29.5 |
| BMI data available | 653 | 34.9 |
| Hypertension data available | 730 | 39.0 |
| Hyperlipidemia data available | 634 | 33.8 |
| Diabetes data available | 634 | 33.8 |

Supplementary Table 2. Distribution of vascular risk score and individual vascular/lifestyle-related factors in the study cohort

| **Variable** | **Category** | **N or value** | **%** |
| --- | --- | --- | --- |
| VRS (complete cases only) | Mean ± SD | 1.16 ± 1.05 | — |
|  | 0 | 169 | 30.6 |
|  | 1 | 204 | 37.0 |
|  | 2 | 116 | 21.0 |
|  | 3 | 47 | 8.5 |
|  | 4 | 16 | 2.9 |
| BMI > 25 | No | 477 | 73.0 |
|  | Yes | 176 | 27.0 |
| Hypertension | No | 512 | 70.1 |
|  | Yes | 218 | 29.9 |
| Hyperlipidemia | No | 381 | 60.1 |
|  | Yes | 253 | 39.9 |
| Diabetes | No | 547 | 86.3 |
|  | Yes | 87 | 13.7 |
| Smoking | No | 390 | 83.0 |
|  | Yes | 80 | 17.0 |
| Alcohol | No | 321 | 68.3 |
|  | Yes | 149 | 31.7 |

Supplementary Table 3. Age and sex distribution of participants across disease groups

| **Diagnosis** | **N** | **Age, years** | **Female, n (%)** |
| --- | --- | --- | --- |
| Autoimmune | 206 | 41.8 ± 13.5 | 151 (73.3%) |
| Tumor | 237 | 47.4 ± 15.2 | 99 (41.8%) |
| Dementia | 80 | 71.4 ± 7.1 | 52 (65.0%) |
| MCI | 159 | 65.5 ± 7.6 | 105 (66.0%) |
| OSA | 51 | 9.0 ± 1.1 | 21 (41.2%) |
| PBC | 27 | 51.0 ± 12.3 | 17 (63.0%) |
| PNE | 37 | 10.1 ± 1.2 | 16 (43.2%) |
| PWML | 19 | 0.7 ± 0.1 | 8 (42.1%) |
| NT1 | 35 | 28.3 ± 12.5 | 16 (45.7%) |
| ALS | 34 | 55.7 ± 10.9 | 9 (26.5%) |
| Overall | 885 | 46.4 ± 21.4 | 494 (55.8%) |

Supplementary Table 4. Concordance between the main normative OEF model and sensitivity-derived trajectories

| **Metric** | **Sensitivity analysis** | **Pearson’s r** | **P value** | **RMSE** |
| --- | --- | --- | --- | --- |
| Median trajectory | Stricter QC | ≈1.00 | <0.0001 | 0.00090 |
|  | Balanced resampling | ≈1.00 | <0.0001 | 0.0074 |
|  | Split-half | ≈1.00 | <0.0001 | 0.0034 |
|  | Bootstrap | ≈1.00 | <0.0001 | 0.0023 |
|  | LOSO | ≈1.00 | <0.0001 | 0.0015 |
| Growth rate | Stricter QC | ≈1.00 | <0.0001 | 5.24 × 10^-5^ |
|  | Balanced resampling | 0.98 | <0.0001 | 0.00022 |
|  | Split-half | ≈1.00 | <0.0001 | 7.95 × 10^-5^ |
|  | Bootstrap | ≈1.00 | <0.0001 | 0.00020 |
|  | LOSO | ≈1.00 | <0.0001 | 2.38 × 10^-5^ |

Note: Predicted median OEF values and corresponding growth rates were sampled at 1-year intervals across the lifespan. Concordance with the main model was quantified using Pearson’s correlation coefficient and RMSE. QC, quality control; LOSO, leave-one-site-out.

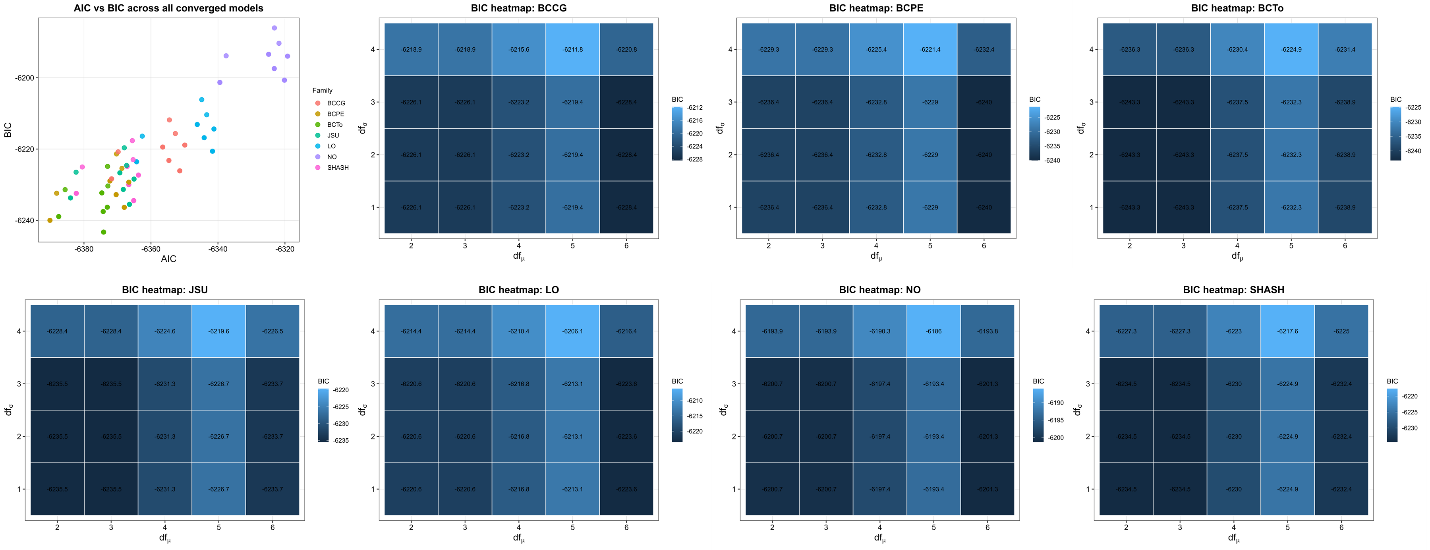

Supplementary Figure S1: Comparison of Akaike Information Criterion (AIC) and Bayesian Information Criterion (BIC) and AIC across different distribution family and model complexity in GAMLSS fitting.

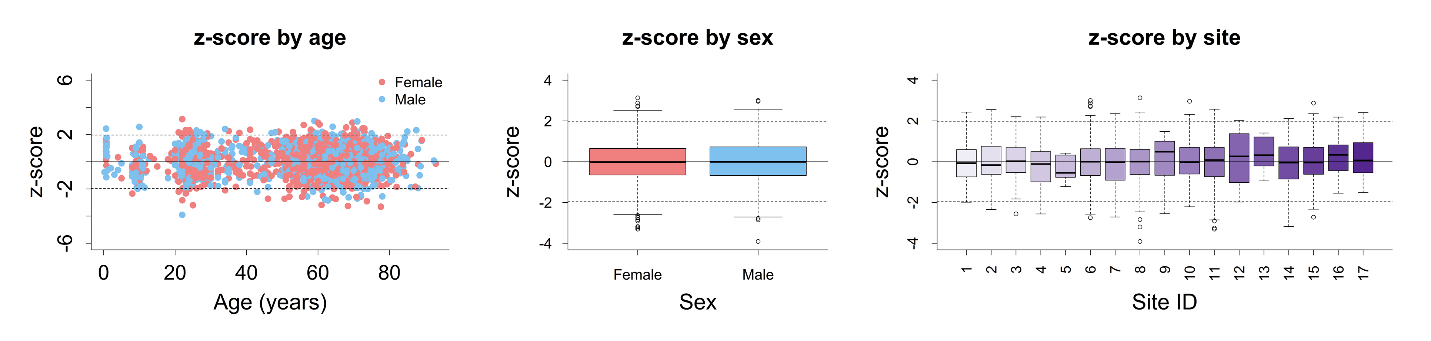

Supplementary Figure S2: Distribution of individual OEF z-scores by age, by sex and across different sites.

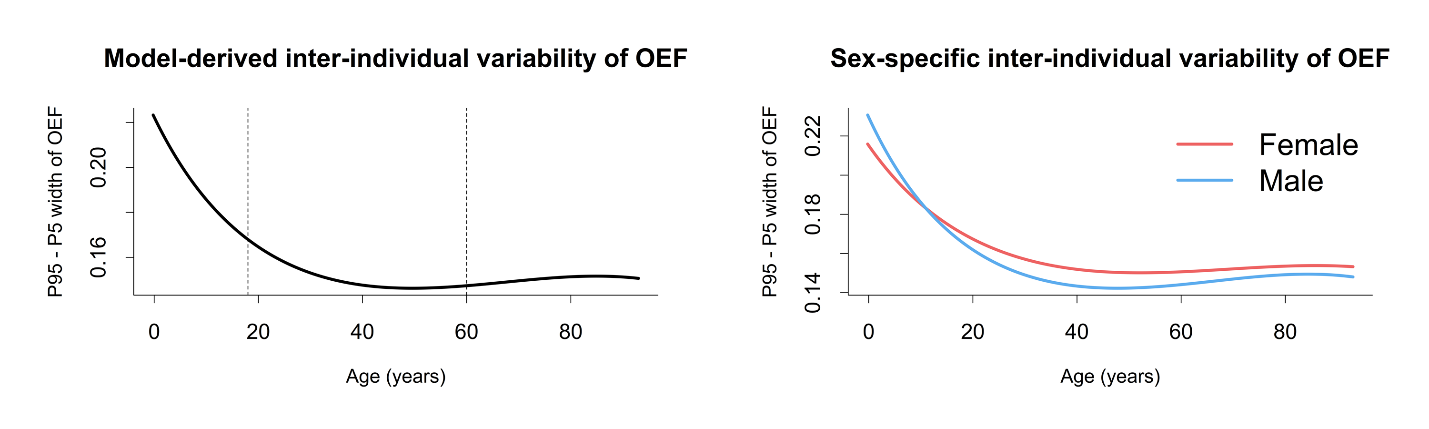

Supplementary Figure S3: Inter-individual variability of OEF as a function of age.

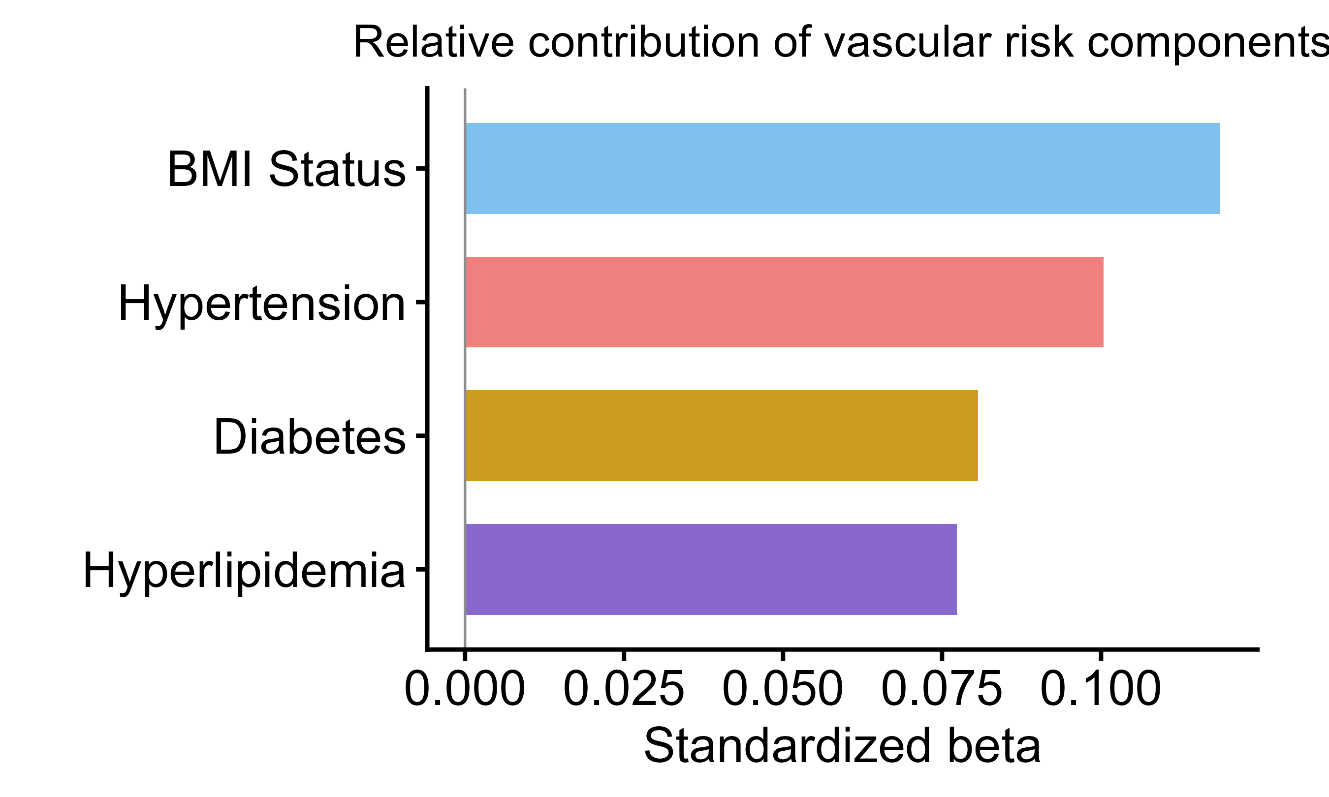

Supplementary Figure S4: Relative contribution of each vascular risk factor to individual OEF deviation from multivariable model.
